# COVID19-Tracker: A shiny app to perform comprehensive data visualisation for SARS-CoV-2 epidemic in Spain

**DOI:** 10.1101/2020.04.01.20049684

**Authors:** Aurelio Tobías, Joan Valls, Pau Satorra, Cristian Tebé

## Abstract

Data visualization is an essential tool for exploring and communicating findings in medical research, especially in epidemiological surveillance. The COVID19-Tracker web application systematically produces daily updated data visualization and analysis of the SARS-CoV-2 epidemic in Spain. It collects automatically daily data on COVID-19 diagnosed cases, and mortality from February 24th, 2020 onwards. Several analyses have been developed to visualize data trends and estimating short-term projections; to estimate the case fatality rate; to assess the effect of the lockdown measures on the trends of incident data; to estimate infection time and the basic reproduction number; and to analyse the excess of mortality. The application may help for a better understanding of the SARS-CoV-2 epidemic data in Spain.

## 1. INTRODUCTION

The first confirmed cases of SARS-CoV-2 in Spain were identified in late February 2020 (1). Since then, Spain became, by the April 27^h^, the second most affected country worldwide (236,199 diagnosed cases) and recorded the third number of deaths (23,521 deaths) due to the SARS-CoV-2 pandemic (2). Since March 16^th^, lockdown measures oriented on flattening the epidemic curve were in place in Spain, restricting social contact, reducing public transport, and closing businesses, except for those essential to the country’s supply chains (3). However, these measures, even they were in the right direction, did not show to be enough to change the rising trend of the epidemic. For this reason, a more restrictive lockdown was suggested (4), and eventually undertaken by the Spanish Government on March 30^th^ (5). On April 13th, a partial opening of the economic activity was allowed by the Spanish Government, allowing non essential activities from businesses not open to public.

Data visualization and analysis is an essential tool for exploring and communicating findings in medical research, and especially in epidemiological surveillance. It can help researchers and policymakers to identify and understand trends that could be overlooked if the data were reviewed in tabular form. We have developed a Shiny app that allows users to evaluate daily time-series data from a statistical standpoint. The COVID19-Tracker app systematically produces daily updated data visualization and analysis of SARS-CoV-2 epidemic data in Spain. It is easy to use and fills a role in the tool space for visualization, analysis, and exploration of epidemiological data during this particular scenario.

## 2. SOFTWARE AVAILABILITY AND REQUIREMENTS

The COVID19-Tracker app has been developed in RStudio (6), version 1.2.5033, using the Shiny package, version 1.4.0. Shiny offers the ability to develop a graphical user interface (GUI) that can be run locally or deployed online. Last is particularly beneficial to show and communicate updated findings to a broad audience. All the analyses have been carried out using R, version 3.6.3.

The application has a friendly structure based on menus to shown data visualization for each of the analyses currently implemented: projections, fatality rates, intervention, infection time, reproducibility number and mortality register analysis (Figure 1).

**Figure 1.**
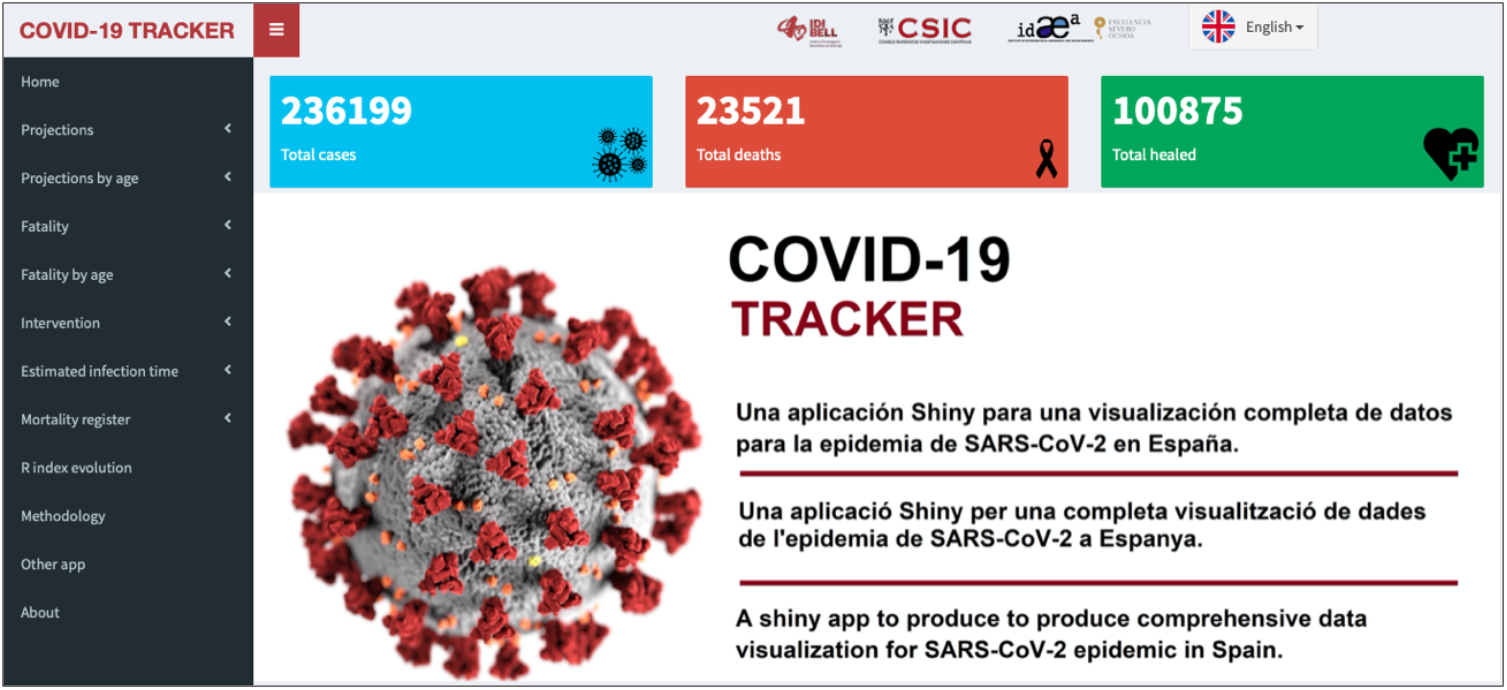
Home page of the COVID19-Tracker application, for visualization and analysis of data from the SARS-CoV-2 epidemic in Spain. Available at: https://ubidi.shinyapps.io/covid19/

– ***Projections*** and ***Projections by age*** display the trends for diagnosed cases and mortality since the epidemic began, and estimates a 3-day projection.
– ***Fatality*** and ***Fatality by age*** display the trends for the case fatality rates.
– **Intervention** calculates and displays the effect of the lockdown periods on the trend of incident daily diagnosed cases and mortality.
– **Infection time** estimates and displays the incubation period for COVID-19 between the interval of exposure to SARS-CoV-2 and the date of COVID-19 diagnosis.
– **Reproducibility number** estimates and displays the average number of secondary cases of disease caused by a single infected individual over his or her infectious period.
– **Mortality registry** displays the evolution of the observed all-cause mortality during the epidemic. It also compares the observed and expected deaths to assess the excess of mortality.

Two additional menus to describe the ***Methodology***, to describe the statistical details on the analyses already implemented, and ***Other apps***, which collects applications also developed in Shiny by other users to follow the COVID19 epidemic in Spain and globally.

The app has an automated process to update data and all analyses every time a user connects to the app. It is available online at the following link: https://ubidi.shinyapps.io/covid19/ and shortly free available on github as an R package. The displayed graphs are mouse-sensitive, showing the observed and expected number of events through the plot. Likewise, when selecting any plot, the application allows the option of downloading it as a *portable network graphic* (* .png). All menus are available in English, Spanish, and Catalan.

## 3. DATA SOURCES

We collected daily data on COVID-19 diagnosed cases and mortality, from February 24^>th^ onwards. Data is collected automatically every day daily from the Datadista Github repository (7). This repository updates data according to the calendar and rate of publication of the Spanish Ministry of Health/Instituto de Salud Carlos III (8).

## 4. METHODS

### 4.1. Projections

To estimate the observed data trends for the number of events, we used a Poisson regression model (9), allowing for over-dispersion (10), fitting lineal, quadratic and cubic terms:

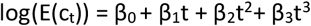

Where *t* = 1, 2, …, T, represents the time unit (from the first observed day until the last, T consecutive days in total), and *c*_*t*_ is the number of events. The estimated regression parameters and their standard errors are used to obtain the short-term projections, up to three days, and their 95% confidence interval (95% CI).

Results are available nationwide by default, and at the regional level accessing to the dropdown menu for this purpose (Figure 2a). Trends and projections are also calculated by age group (0-39, 40-49, 50-59, 60-69, 70-79, and 80 or more years) (Figure 2b).

**Figure 2.**
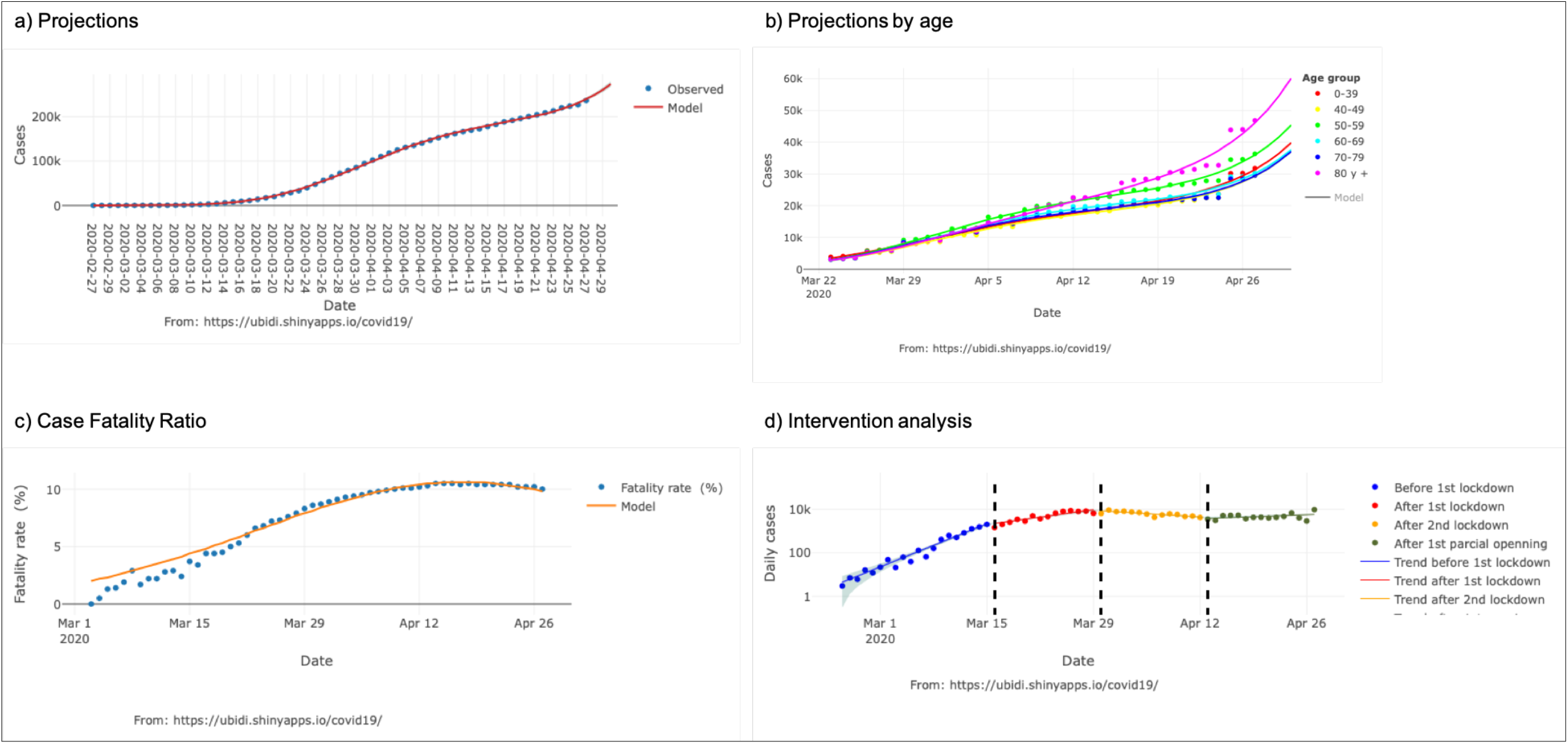
Standard output display of the COVID19-Tracker application (results updated to April 27^th^, 2020), for trend analysis and its 3-day projection at the national level (a) and by age group (b), of the fatality rate (c), and intervention analysis to evaluate the effect of alarm states on incident data (d).

We should note that in previous versions of the application, linear or quadratic models were considered. Based on the evolution of the epidemic, these models were compared using a likelihood ratio test based on their deviances. In the current version, the cubic model is the one showing the best goodness of fit. In any case, the models are regularly being evaluated, in case a model reformulation with a better fit is necessary during the course of the epidemic.

### 4.2. Case fatality rate

The case fatality rate is defined as the ratio between the number of deaths and the diagnosed cases (11). Thus, an offset is fitted into the Poisson regression model, also allowing for overdispension, as the logarithm of the diagnosed cases:

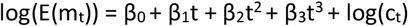

Where m_t_ is the daily number of deaths, and c_t_ is the daily number of diagnosed cases. Case fatality rates are also calculated for the same age groups (Figure 2c).

We should acknowledge that it is not possible to make an accurate estimate of the case fatality rates due to underreporting of cases diagnosed in official statistics (12). Nonetheless, the estimation and monitoring of the case fatality rates monitoring are of espeical interest in the current epidemic scenario.

### 4.3. Intervention analysis

To assess the effect of the lockdown on the trend of incident diagnosed cases and mortality, we used an interrupted time-series design (13). The data is analyzed with quasi-Poisson regression with an interaction model to estimate the change in trend:

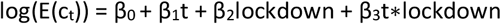

Where *lockdown* is a variable that identifies the intervals before and during the lockdown periods imposed by the Spanish Government (3, 5) (0=before March 15^th^,

2020; 1=between March 16^th^ and March 29^th^, 2020; 2= between Mach 30^th^ and April 12^th^, 2020; 3=after April 13^th^, 2020).

We should acknowledge that this is a descriptive analysis without predictive purposes (Figure 2d). For an easy interpretation, and comparison of the effectiveness of lockdown measures between countries, a linear trend is assumed before and during each lockdown period (14). Although not accounted for residual autocorrelation, the estimates are unbiased but possibly inefficient. This analysis also shows the results nationwide in table reporting the daily percentage (%) mean increase, and its 95% CI.

### 4.4. Estimation of time to infection

Lauer et al. (15) have recently analyzed the incubation period for COVID-19 in a cohort of symptomatic patients. From each patient, they collected the interval of exposure to SARS-CoV-2 and the date of appearance of symptoms. They assumed that the incubation time would follow, as in other viral respiratory tract infections, a Lognormal distribution.

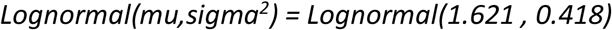

In a naïve exercise with some limitations, we have replicated this distribution in the group of diagnosed cases to approximate the date of exposure to SARS-CoV-2 recursively:

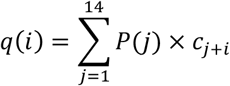

Where p is the number of diagnosed cases on day i, q is the number of infected cases on day i-j, and j = 1, 2, …, 14 the maximum time it is expected that the disease can develop. P(j) is the probability of presenting symptoms on day j according to a Lognormal law with the parameters defined by Lauer et al. (15)

To estimate the last 14 days, since the information on the diagnosed cases was not available for the next 14 days, a quadratic model was used to project diagnosed cases. These latest estimates are displayed in the application with a different color.

Results are available nationwide by default (Figure 3a), and at the regional level accessing to the dropdown menu for this purpose.

**Figure 3.**
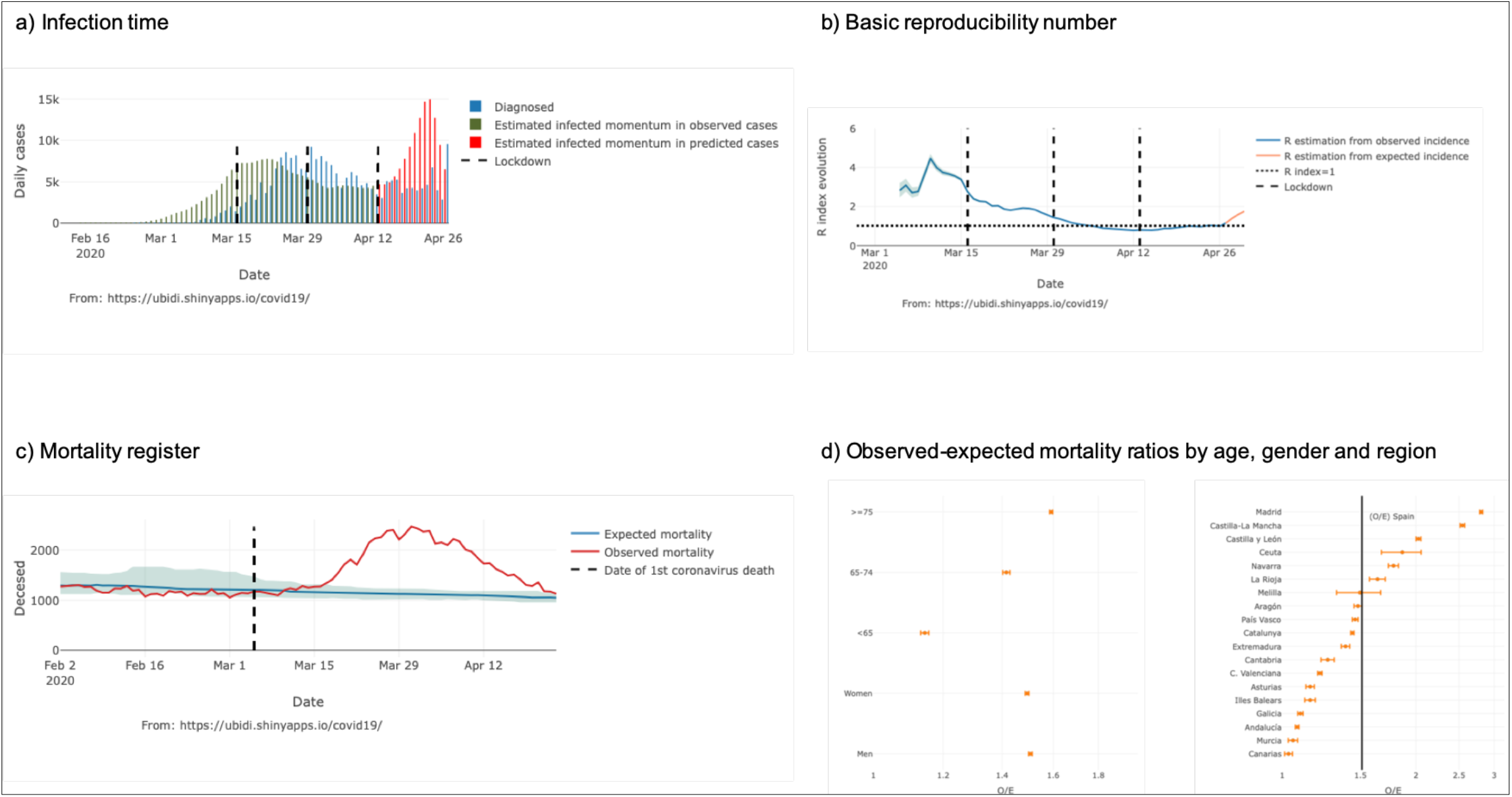
Standard output display of the COVID19-Tracker application (results updated to April 27^th^, 2020), estimated infected time (a) and basic reporicibility number (b), mortality register (c), and observed versus expected mortality ratios by age, gender and region (d).

### 4.5. Basic reproduction number

The basic reproduction number (R_0_) is the average number of secondary cases of disease caused by a single infected individual over his or her infectious period (16). This statistic, which is time and situation specific, is commonly used to characterize pathogen transmissibility during an epidemic. The monitoring of R_0_ over time provides feedback on the effectiveness of interventions and on the need to intensify control efforts, given that the goal of control efforts is to reduce R_0_ below the threshold value of 1 and as close to 0 as possible, thus bringing an epidemic under control.

Here, we use the R package *EpiEstim* to estimate the basic reproduction number R_t_ using the Wallinga and Teunis method (16). This parametric method assumes a gamma distribution for the serial interval. The serial interval is the time between the onset of symptoms in a primary case and the onset of symptoms of secondary cases, which is needed to estimated R_t_ over the course of an epidemic.

The mean and standard deviation of the serial interval distribution can vary depending on the disease (16). Recently Nishiura et al. (17) estimated a mean and standard deviation for COVID-19 of 4.7 and 2.9 days, being theses the values we are using in our analysis application foer the gamma a priori distribution.

Results are available nationwide by default (Figure 3b), and at the regional level accessing to the dropdown menu for this purpose.

### 4.6. Mortality registry

We show the evolution of the observed all-cause mortality during the epidemic, jointly with the expected mortality according to MoMo the model by the Instituto de Salud Carlos III (18).

Results are available nationwide by default, and at the regional level accessing to the dropdown menu for this purpose (Figure 3c). We also calculated the observed and expected mortality ratios, and their 95% CI, by age, gender and region, since March 4^th^, 2020, the date on which the first fatality from COVID-19 is official reported in Spain (7, 8) (Figure 3d)

### 5. Further developing

So far, the COVID19-Tracker app has been very well received online, with a large number of connections generating an outsized memory usage on our server (Figure 4).

**Figure 4.**
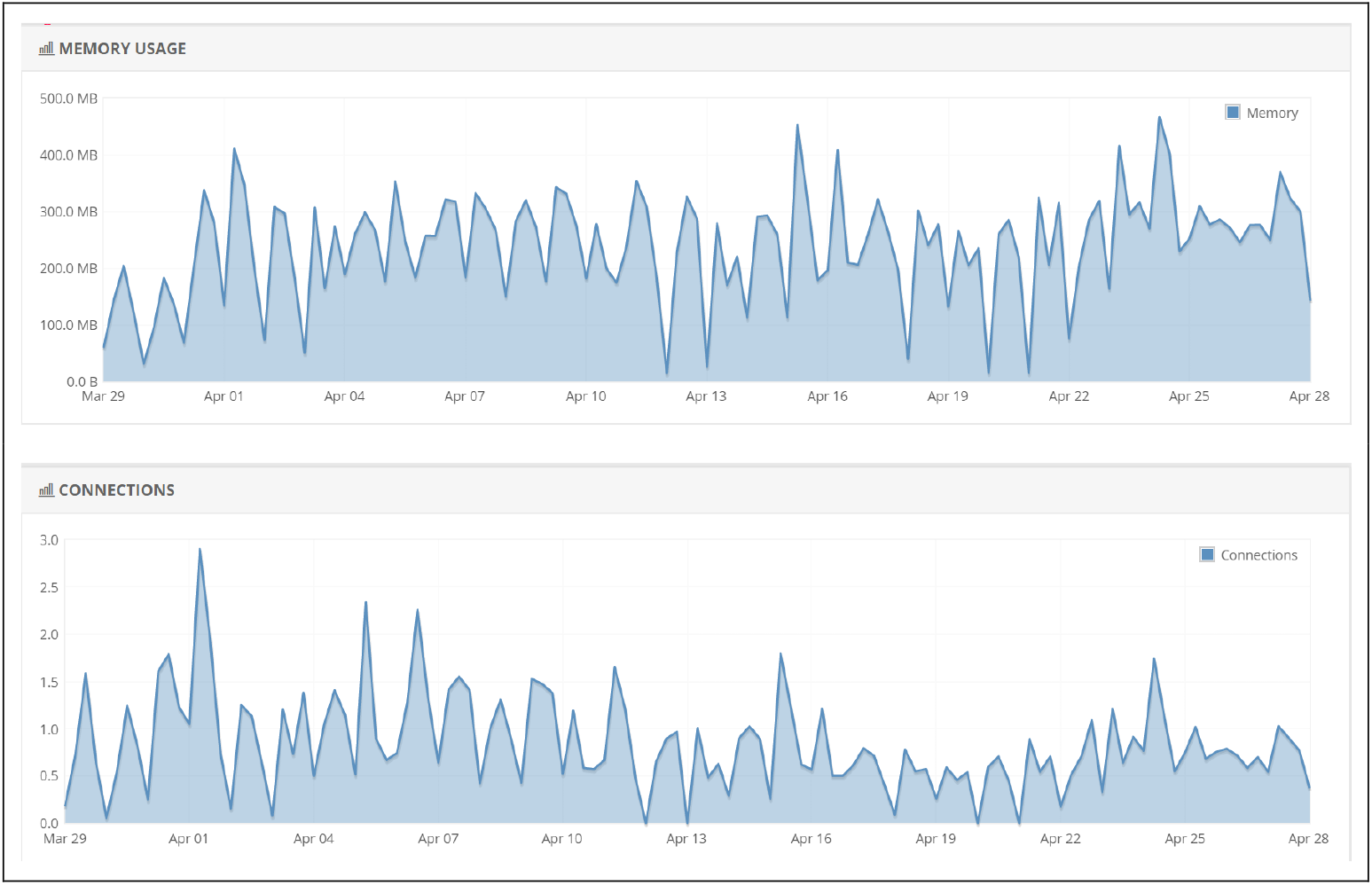
Number of connections and memory usage since March 27^th^ to April 27^th^, 2020.

We keep improving the application by uploading new data visualizations, which may help for a better understanding of the SARS-CoV-2 epidemic data in Spain. Moreover, the COVID19-Tracker app could also be extensible to data visualizations across other countries and geographical regions.

## Discussion

The COVID19-Tracker application presents a set of tools for updated analysis and graphic visualization that can be very useful for a better understanding of the evolution of the COVID-19 epidemic in Spain and its epidemiological surveillance.

As limitations, we should be note that the application does not take into account the changes in the definition of a case diagnosed by COVID-19, nor the population exposed. So, the number of events is modeled directly instead of the incidence rate, assuming that the entire population is at risk, except for the case fatality rate. On the other hand, the analyzes are not free from the biases linked to the source of information provided by the Ministry of Health (8), being collected on a daily basis through the Datadista github (7).

We continue to plan improvements to the app to include new analytics and visualizations. Also, the application could be extensible for use in other countries or geographic areas. In summary, this application, easy to use, come to fill a gap in this particular scenario for the visualization of epidemiological data for the COVID-19 epidemic in Spain.

## Data Availability

Data on COVID-19 diagnosed cases, intensive care unit (ICU) admissions and mortality is available from the Datadista github repository. This repository updates data according to the calendar and rate of publication of the Spanish Ministry of Health/Instituto de Salud Carlos III.

https://github.com/datadista/datasets/tree/master/COVID%2019

https://covid19.isciii.es/

## Funding

None.

### Acknowledgements

None.

